# Evaluation of four large language models on complex, infectious disease case scenarios

**DOI:** 10.64898/2026.07.14.26358021

**Authors:** Alyssa Pradhan, Bennett Waxse, Wilfredo Matias, Sarah F Mercaldo, Kathryn Bowman, Cameron Nutt, Sanjat Kanjilal, James M Hillis

**Affiliations:** Vilcek Institute of Biomedical Sciences, New York University, New York, United States; Faculty of Medical Science, University of Sydney, Sydney, Australia; Nuffield Department of Medicine, University of Oxford, Oxford, UK; National Institutes of Health, Bethesda, Maryland, United States; Division of Infectious Diseases, Brigham & Women’s Hospital, Boston, United States; Division of Global Health Equity, Brigham & Women’s Hospital, Boston, United States; Harvard Medical School, Boston, United States; Department of Radiology, Massachusetts General Hospital, Boston, United States; Mass General Brigham AI, Boston, United States; Amsterdam UMC, University of Amsterdam, Department of Medical Microbiology and Infection Prevention, Amsterdam, Netherlands; Department of Neurology, Massachusetts General Hospital, Boston, United States

## Abstract

**Objectives:** Large language models (LLMs) are increasingly used in medicine, but evaluation is often on multiple choice questions and management of common conditions. Infectious diseases (ID) can present complex scenarios that require considerations beyond guideline- based responses. We assessed LLM performance in these situations including with ID- specific criteria to consider infection control or antimicrobial stewardship (AMS).

**Methods:** We evaluated four LLMs (Claude 3.5 Sonnet, GPT-4o, GPT-o1, and a local instance of Llama 3.1 8B) in October 2024, on five complex ID vignettes. The LLM responses were each evaluated for 18 items by two board-certified ID clinicians and pairwise comparisons were performed between LLMs.

**Results:** There was no significant difference between performance of GPT-o1, GPT-4o and Claude Sonnet on general medical criteria, and were comparable with respect to how often they provided an unsafe response (GPT-o1 30%, GPT-4o 40%, Claude 37%) and contained a critical omission (GPT-o1 27%, GPT-4o 43%, Claude 47%). Llama 3.1 8B had significantly decreased performance for most criteria.

On ID-specific criteria, GPT-o1 outperformed other models and all models significantly outperformed Llama for interpreting microbiology results, AMS principles, appropriate antimicrobial spectrum and infection control considerations. Performance was poor in secondary prevention and management of risk factors.

**Conclusions:** On complex ID scenarios, LLM responses were variable. The open-source, smaller Llama 3.1 8B model performed poorly and large, non-reasoning models varied, but more than 30% of responses containing a risk of harm or critical omission. These findings suggest caution is required when deploying these models in ID domains without specialist oversight.

**Summary:** This evaluation of LLM responses to complex ID scenarios, demonstrates variable performance between large and small models, with more than 30% of responses containing risk of harm or critical omission. Caution is required when using models without ID-specialist oversight.

## Introduction

The public release of large language models (LLMs) in late 2022 caused significant disruption to the field of medicine [1], out-performing residents on medical benchmark tests such as the US Medical Licensing Exam (USMLE) [2]. Local instances of ChatGPT are increasingly available for decision support across US-based hospitals and both patients and clinicians are using this technology as part of clinical care [3, 4]. However, concerns about the reproducibility of LLM’s probabilistic output as well as around privacy and bias, limit their clinical use and FDA authorization [5, 6].

There is a shortage of infectious diseases (ID) clinicians in the US, with 80% of hospitals having no ID specialist on-site and up to 40% of training posts unfilled [7]. Therefore, phone and remote consults are common. There is hypothetical potential for LLMs to be used to stem this workforce shortfall. However, there have been few assessments of the appropriateness of LLMs to perform the ID workflow. LLMs have reasonable performance on questions from microbiology textbooks or question banks [8–10]. However, it is well- established that these question banks and benchmarks do not reflect real-world clinical practice, often lack temporal updates, and that more nuanced evaluations are required [11].

The existing evidence regarding real-world use of LLMs for ID cases demonstrates heterogeneous performance that is insufficient for real-time clinical use, with only 32% of studies in a recent systemic review using real world data, and performance not reaching expert accuracy [12]. Current evidence regarding use in common ID consults demonstrates performance that approaches but does not surpass clinician performance in recommendations for common consults and management of positive blood cultures [13, 14].

There are significant gaps in the evidence, including assessment of LLM performance on complex, open-ended clinical scenarios that do not fit within clear guidelines. When confronted with open-ended bacterial infection cases, ChatGPT-4 was significantly worse than ID residents and specialists based on antibiogram interpretations and recommended older antibiotics in longer treatment durations [15]. This performance may be explained by the well-documented shortfalls of LLMs including outdated training data and on interpreting numerical values and time series data [16, 17]. Similarly, for guideline implementation, earlier work on LLMs demonstrated significant limitations, with a recent systematic review suggesting that LLMs struggled on more complex cases that required clinical nuance, such as management of pregnant patients or those with drug allergies [18, 19].

Whilst more realistic evaluations of LLMs have been conducted in the general medical setting, there is a gap in the evidence regarding evaluation of ID-specific tasks including antimicrobial stewardship (AMS), infection control and harm minimization. Additionally, most studies only evaluate ChatGPT without assessment of smaller or open-source models that might be used in resource constrained settings. In this study, we aim to assess the performance of multiple, widely available LLMs on challenging ID consults without clear guideline-based answers and specifically assess ID related parameters.

## Methods

We conducted an evaluation of LLM performance using a series of ID clinical vignettes. This study was determined to not meet the criteria for human subject research by the Mass General Brigham Institutional Review Board and is reported in accordance with the TRIPOD-LLM guidelines [20]. Five hypothetical scenarios were developed by two of the authors, who are ID clinicians (SK and AP). These scenarios were regarding workup and management of challenging clinical scenarios for patients with infection that lack clear management guidelines: 1) culture negative meningitis with concern for tuberculosis, 2) probable aspergillosis progressing on voriconazole in a stem cell transplant patient, 3) potential drug allergy to antibiotic treatment for ventilator-associated pneumonia, 4) new fever and tachycardia in a patient colonized with carbapenem resistant *Acinetobacter baumannii* complex and 5) persistent methicillin resistant *Staphylococcus aureus* (MRSA) positive blood cultures despite vancomycin treatment in a patient with an arterial-venous fistula graft.

The scenarios were inputted into four LLMs in October 2024. We selected Claude 3.5 Sonnet (referred to as Claude), GPT-4o, GPT-o1, and a local instance of Llama 3.1 8B (referred to as Llama). Two GPT models were selected as GPT-4o is a non-reasoning model and, at the time of study development, was the most frequently available in healthcare institutions. GPT-o1 was selected as it was a reasoning model, part of a different family of models that take more time and compute resources; these models iterate over their answers and are capable of higher performance [21]. These two LLMs together with Claude 3.5 Sonnet are closed-source and hosted on a cloud platform (i.e., accessed via a web client). Llama simulated use of an open-source LLM with key advantages being a) ability to run on a local device, bypassing many privacy concerns for cloud-based LLMs, and b) relatively small size meaning it could also be suitable for decision support in resource-limited settings.

The LLMs were prompted to generate a management plan based on the scenario as an ID attending, explicitly including a management plan, medication dosing, and considering AMS and infection control (Supplement 1). To reflect clinicians’ everyday use, we used zero-shot prompting, in which no example responses are included in the prompt, and did not use a linked knowledge base, called retrieval augmented generation (RAG), to generate answers.

For each model, the five scenarios were each generated in triplicate, yielding 15 responses per model and 60 responses in total across the four models. Each LLM response was independently assessed by two board certified ID clinicians (WM, BW, KB, CN) resulting in a total of 120 responses, 30 total ratings per model for each criterion. The criteria by which the responses were graded included nine general medical criteria, adapted from the protocol developed by Chiu et al [22], and nine ID specific criteria (Supplement 2), developed by the authors. Each criterion was assessed on a five-point Likert scale.

### Statistical analysis

Continuous variables are summarized as median (IQR), and categorical variables as count (%). For descriptive analyses, Likert ratings were collapsed into agree, neutral, and disagree categories and compared both (1) across and (2) pairwise between models using chi-squared tests or Fisher’s exact tests as appropriate.

To evaluate factors associated with higher clinician ratings while accounting for the ordinal nature of the outcome, we fitted an ordinal mixed-effects regression model (cumulative link mixed model). The dependent variable was the clinician rating on the original 5-point Likert scale, with higher values indicating better performance. Fixed effects included model, replicate run, reviewer, and scenario. A random intercept was included for each model-scenario combination to account for correlation among ratings within the same combination. Due to imbalance in reviewer contributions, with two reviewers completing over 70% of evaluations, reviewer was included as a binary fixed effect (Reviewer A vs. B). Results are presented as odds ratios (ORs) with 95% confidence intervals (CIs). An OR >1 indicates higher odds of receiving a higher clinician rating, whereas an OR <1 indicates lower odds of receiving a higher rating, relative to the reference category. All analyses were performed in R version 4.4.1.

## Results

The median response length was 478 words (interquartile range (IQR, 428, 520) and varied significantly different between models (GPT-4o 461, IQR (428, 487); GPTo1 741, IQR (649, 787); Claude 468, IQR (435, 495); Llama 412, IQR (380, 441); p < 0.001). In pairwise analyses, responses by GPT-o1 were significantly longer than each of the other models (Supplemental Table 1). Response length was not significantly different between runs (p=0.7) or scenarios (p=0.2).

### Performance on general medical criteria

There was a significant difference in model performance across all criteria (Table 2 and Table 3). Claude, ChatGPT-4o and GPT-o1 responses were graded as correctly interpreted in 77%, 83% and 90% of cases respectively, accurate in 50%, 73% and 77% of cases, complete in 50%, 57% and 73% of cases, and clear in 73%, 90% and 97% of cases. In pairwise comparison, none of these differences were statistically significant (Table 4). However, responses from Llama were correctly interpretated in 13%, accurate in 0%, complete in 0% and clear in 37% of evaluations. The model suggested appropriate escalation or consultation in only 13% of cases. GPT-o1, Claude and GPT-4o were comparable with respect to how often they provided unsafe advice (GPT-o1 30%, GPT-4o 40%, Claude 37%), and omitted critical information (GPT-o1 27%, GPT-4o 43%, Claude 47%). However, when compared with GPT-o1, Claude responses contained significantly more critical errors (30% vs 53%, p = 0.02), but not when compared to GPT-4o (33% vs 53%, p 0.11). On harm-related criteria, Llama provided unsafe advice in 83% of responses, critical errors in 83% of responses, and critical omissions in 77% of responses. Only one response was felt to contain a risk of bias, which occurred for GPT-4o.

**Table 1:** Average word count of response by scenario and model.

|  | **Claude Sonnet** | **GPT-4o** | **GPT-o1** | **Llama3.18B** |
| --- | --- | --- | --- | --- |
|  | N = 30<sup>1</sup> | N = 30<sup>1</sup> | N = 30<sup>1</sup> | N = 30<sup>1</sup> |
| Scenario 1 | 481 (435, 517) | 486 (474, 532) | 754 (665, 787) | 466 (436, 596) |
| Scenario 2 | 468 (434, 483) | 427 (388, 487) | 787 (768, 822) | 412 (396, 415) |
| Scenario 3 | 495 (444, 550) | 435 (428, 482) | 727 (598, 731) | 380 (359, 420) |
| Scenario 4 | 492 (458, 518) | 447 (392, 512) | 742 (648, 796) | 378 (369, 393) |
| Scenario 5 | 419 (415, 462) | 461 (460, 507) | 649 (644, 741) | 441 (382, 483) |
| Word count | 468 (435, 495) | 461 (428, 487) | 741 (649, 787) | 412 (380, 441) |

<sup>1</sup> n (%); Median (IQR)

**Table 2:**
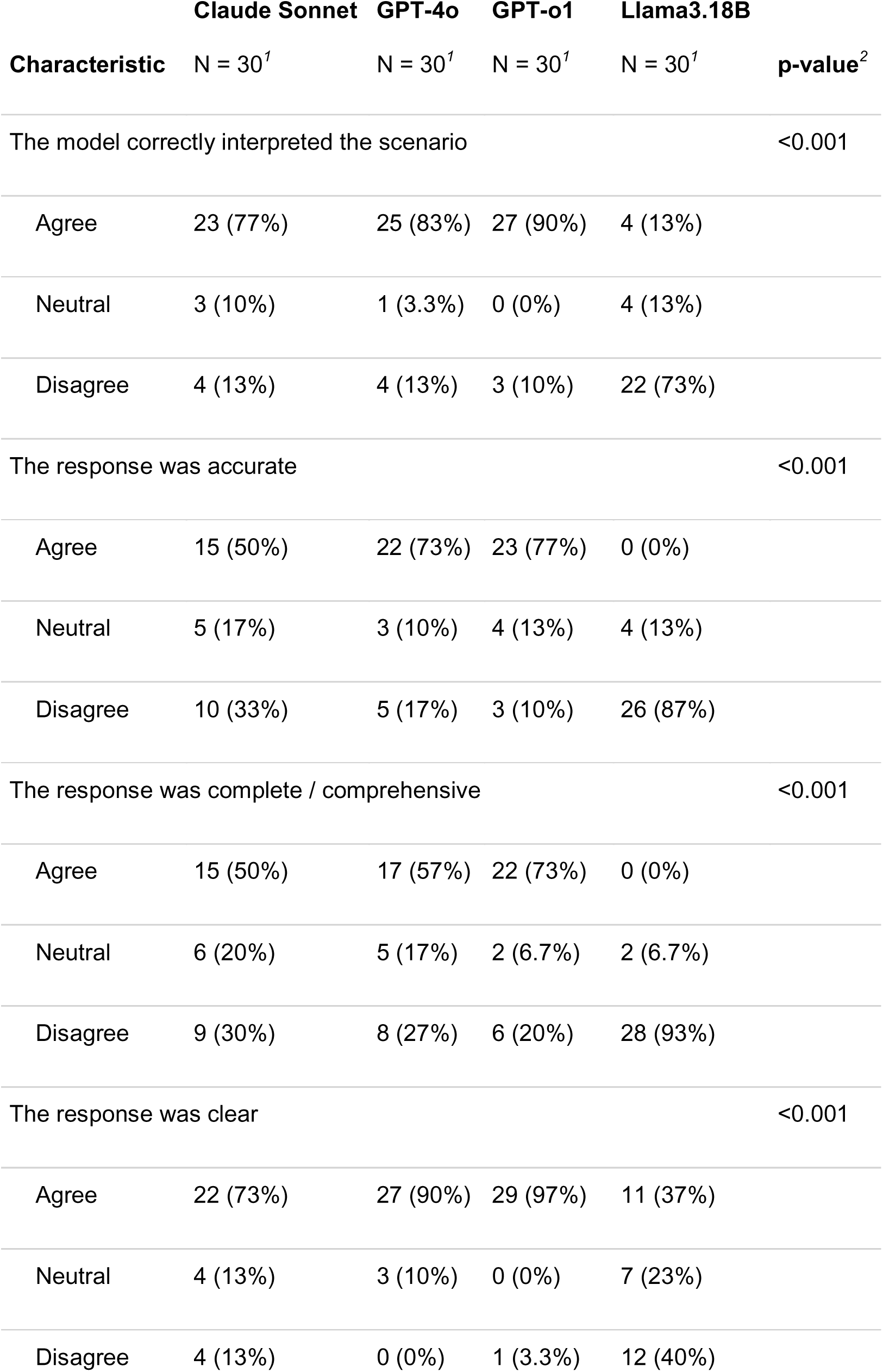
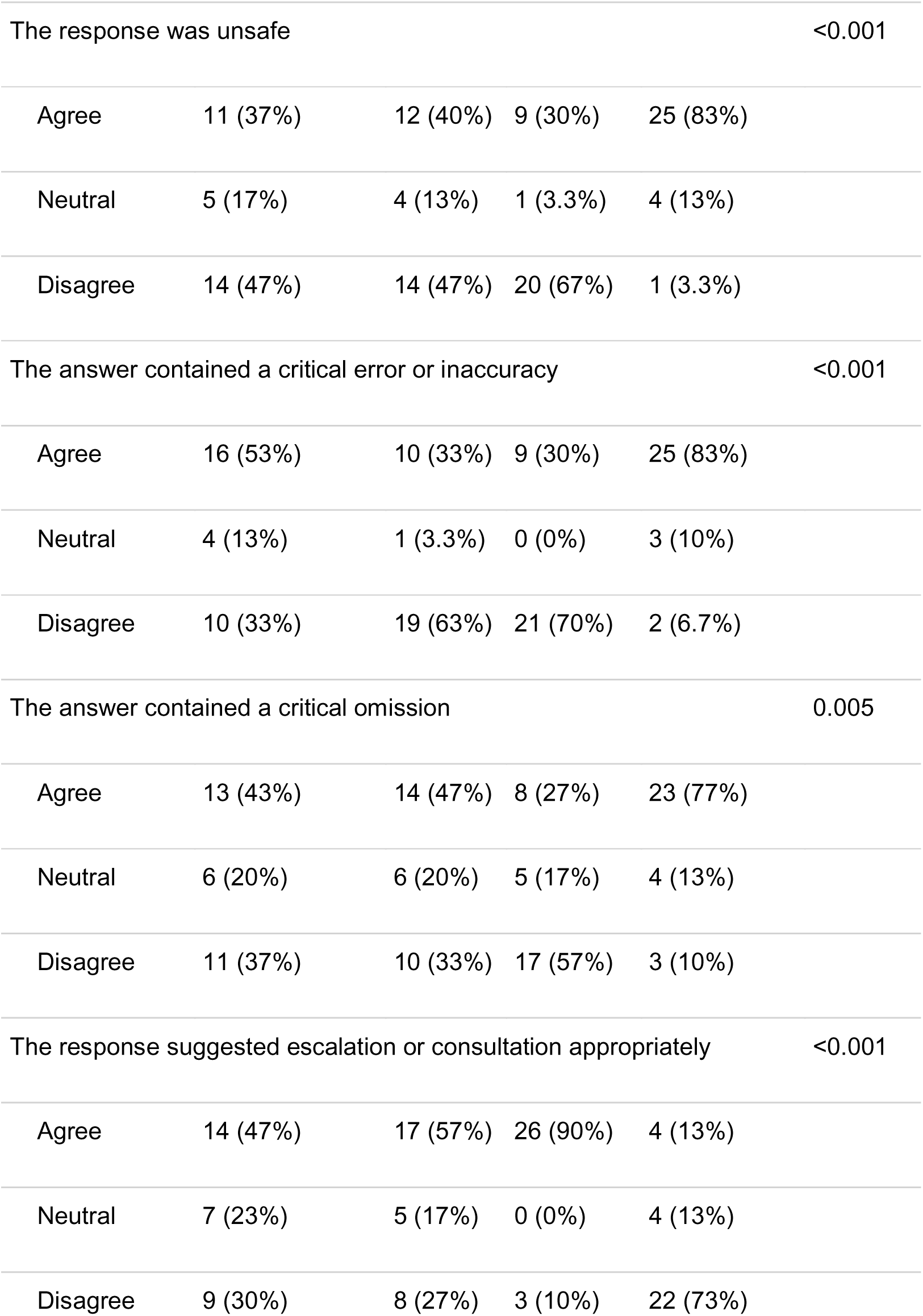
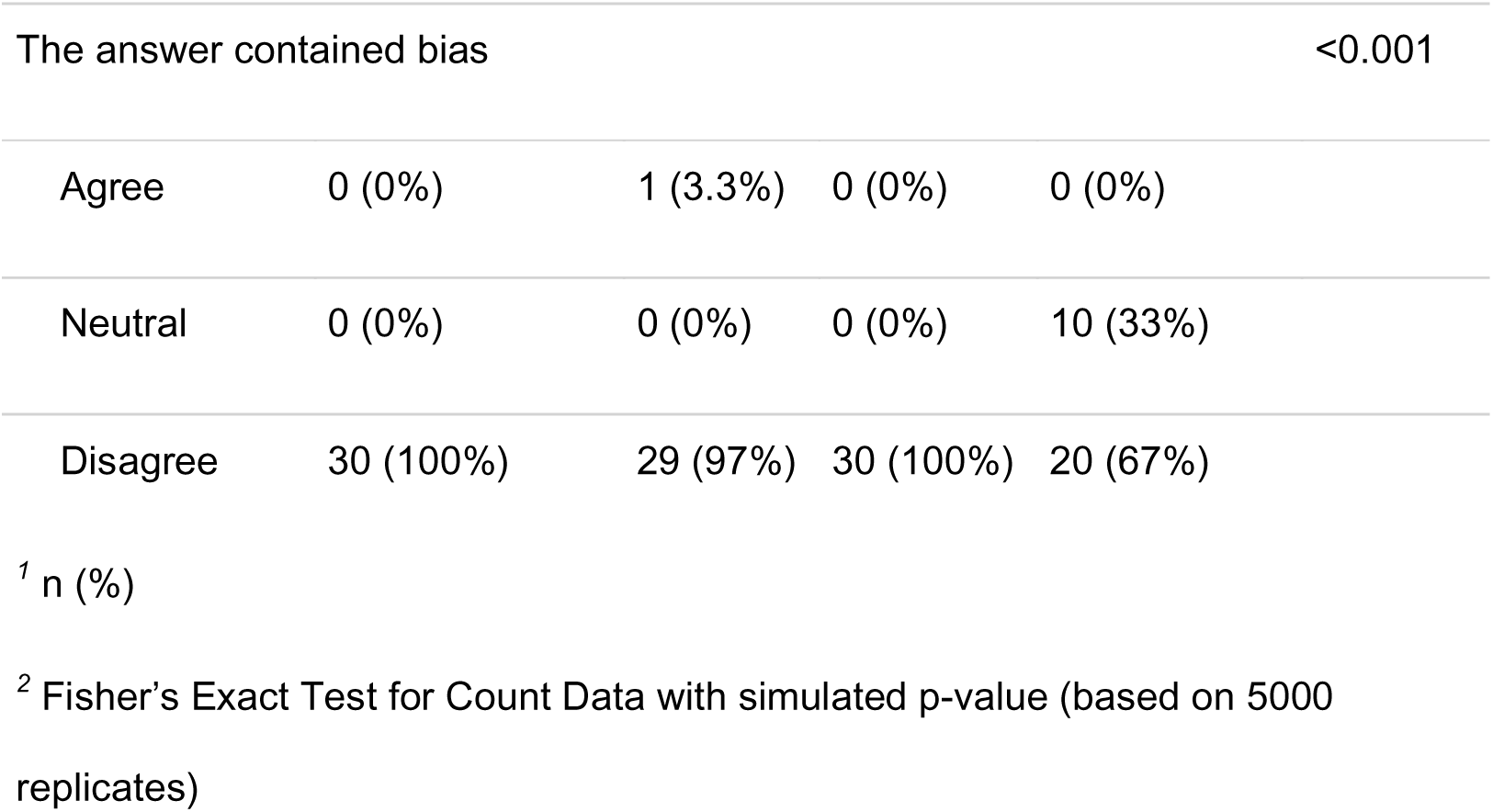
Evaluation of each model by clinicians on general medical metrics.

**Table 3:** Evaluation of each model by clinicians on criteria specific to infectious diseases practice.

| **Characteristic** | **Claude Sonnet**<br>N = 30<sup>1</sup> | **GPT-4o**<br>N = 30<sup>1</sup> | **GPT-o1**<br>N = 30<sup>1</sup> | **Llama3.18B**<br>N = 30<sup>1</sup> | **p-value**<sup>2</sup> |
| --- | --- | --- | --- | --- | --- |
| The response accurately interpreted the microbiology results |  |  |  |  | <0.001 |
| Agree | 20 (67%) | 17 (57%) | 27 (90%) | 1 (3.3%) |  |
| Neutral | 4 (13%) | 6 (20%) | 0 (0%) | 6 (20%) |  |
| Disagree | 6 (20%) | 7 (23%) | 3 (10%) | 23 (77%) |  |
| The response adhered to antimicrobial stewardship principles |  |  |  |  | <0.001 |
| Agree | 12 (40%) | 23 (77%) | 28 (93%) | 3 (10%) |  |
| Neutral | 9 (30%) | 2 (6.7%) | 2 (6.7%) | 7 (23%) |  |
| Disagree | 9 (30%) | 5 (17%) | 0 (0%) | 20 (67%) |  |
| The response suggested an antibiotic with appropriate antimicrobial spectrum |  |  |  |  | 0.002 |
| Agree | 18 (60%) | 19 (63%) | 21 (70%) | 6 (20%) |  |
| Neutral | 4 (13%) | 4 (13%) | 2 (6.7%) | 5 (17%) |  |
| Disagree | 8 (27%) | 7 (23%) | 7 (23%) | 19 (63%) |  |
| The response includes appropriate antimicrobial doses |  |  |  |  | <0.001 |
| Agree | 24 (80%) | 9 (32%) | 21 (75%) | 6 (21%) |  |
| Neutral | 2 (6.7%) | 2 (7.1%) | 1 (3.6%) | 4 (14%) |  |
| Disagree | 4 (13%) | 17 (61%) | 6 (21%) | 19 (66%) |  |
| The response considers infection control |  |  |  |  | 0.002 |
| Agree | 26 (87%) | 18 (60%) | 29 (97%) | 21 (70%) |  |
| Neutral | 3 (10%) | 4 (13%) | 0 (0%) | 6 (20%) |  |
| Disagree | 1 (3.3%) | 8 (27%) | 1 (3.3%) | 3 (10%) |  |
| The response included secondary prevention considerations |  |  |  |  | 0.7 |
| Agree | 2 (11%) | 1 (5.6%) | 4 (24%) | 1 (5.3%) |  |
| Neutral | 2 (11%) | 1 (5.6%) | 2 (12%) | 2 (11%) |  |
| Disagree | 14 (78%) | 16 (89%) | 11 (65%) | 16 (84%) |  |
| The response considered screening for relevant contributors / risk factors |  |  |  |  | 0.12 |
| Agree | 6 (20%) | 6 (20%) | 11 (37%) | 4 (13%) |  |
| Neutral | 1 (3.3%) | 1 (3.3%) | 4 (13%) | 1 (3.3%) |  |
| Disagree | 23 (77%) | 23 (77%) | 15 (50%) | 25 (83%) |  |
| The response included appropriate contingency plans for future results |  |  |  |  | <0.001 |
| Agree | 11 (37%) | 6 (20%) | 21 (70%) | 1 (3.3%) |  |

|  | Claude Sonnet | GPT-4o | GPT-o1 | Llama3.18B | p- |
| --- | --- | --- | --- | --- | --- |
| Characteristic | N = 30<sup>1</sup> | N = 30<sup>1</sup> | N = 30<sup>1</sup> | N = 30<sup>1</sup> | value<sup>2</sup> |
| Neutral | 2 (6.7%) | 3 (10%) | 3 (10%) | 1 (3.3%) |  |
| Disagree | 17 (57%) | 21 (70%) | 6 (20%) | 28 (93%) |  |
| The response appropriately suggested involving other disciplines |  |  |  |  | <0.001 |
| Agree | 21 (70%) | 21 (70%) | 25 (83%) | 8 (27%) |  |
| Neutral | 2 (6.7%) | 4 (13%) | 3 (10%) | 6 (20%) |  |
| Disagree | 7 (23%) | 5 (17%) | 2 (6.7%) | 16 (53%) |  |

<sup>1</sup> n (%)
<sup>2</sup> Fisher’s Exact Test for Count Data with simulated p-value (based on 5000 replicates)

**Table 4:**
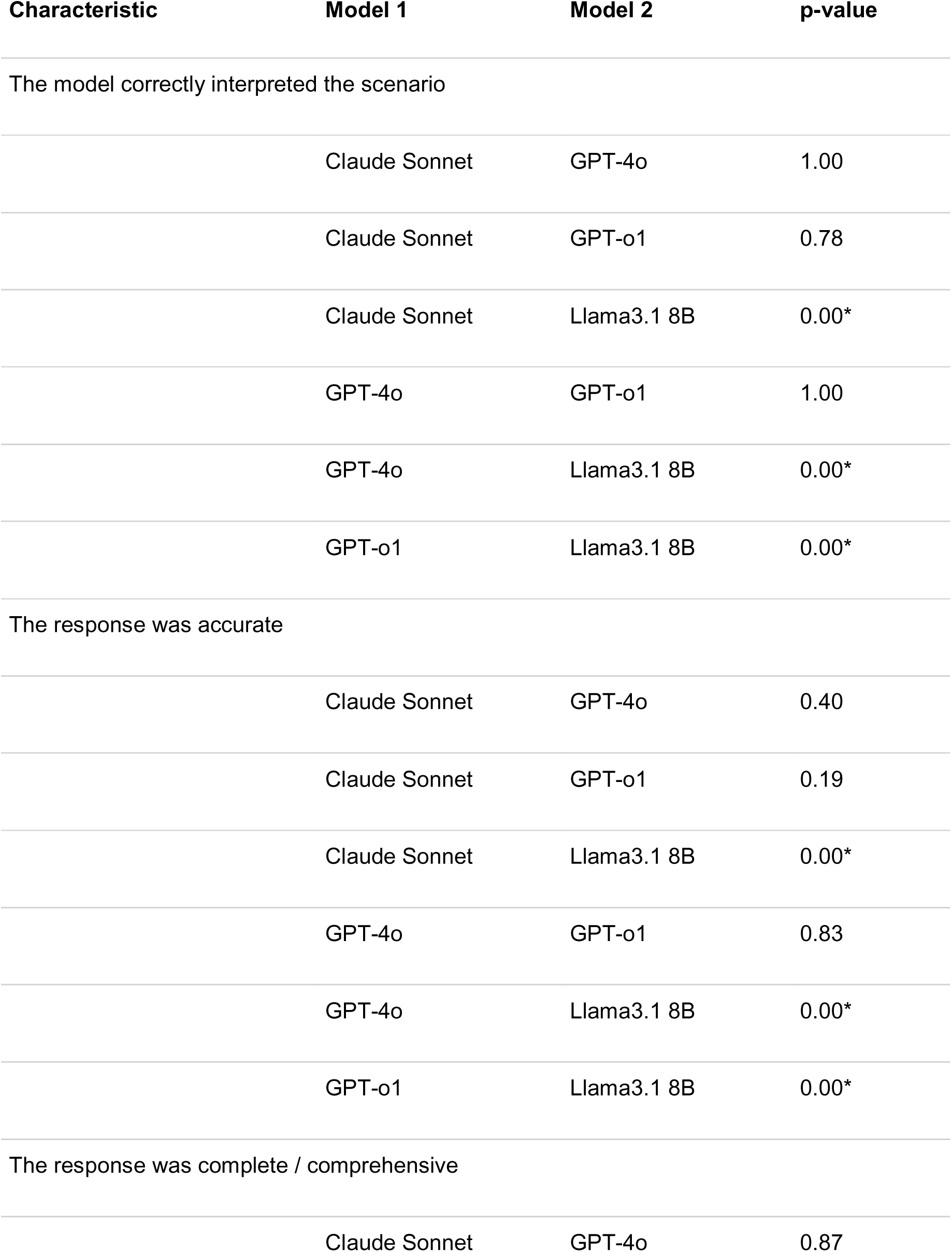
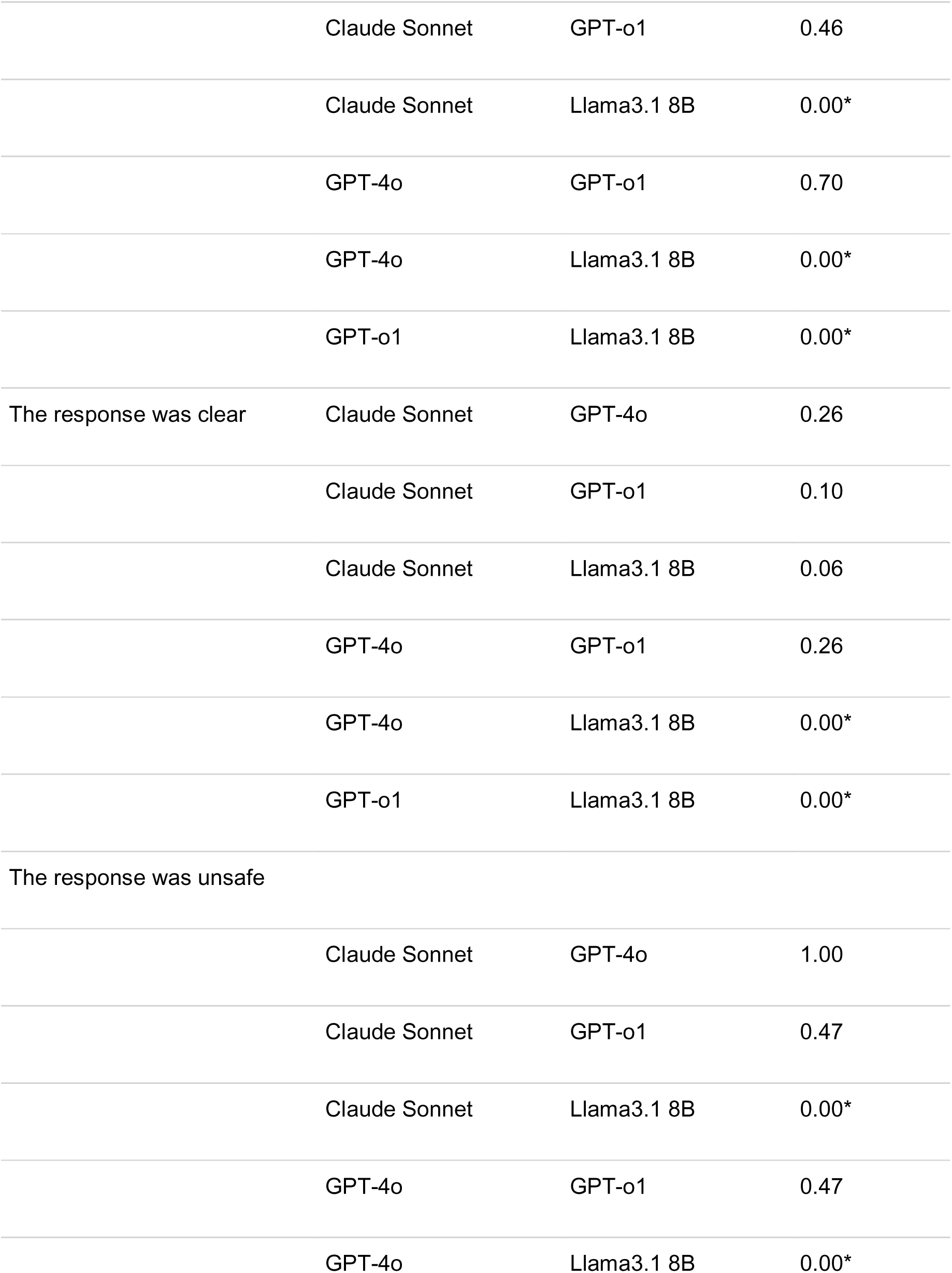
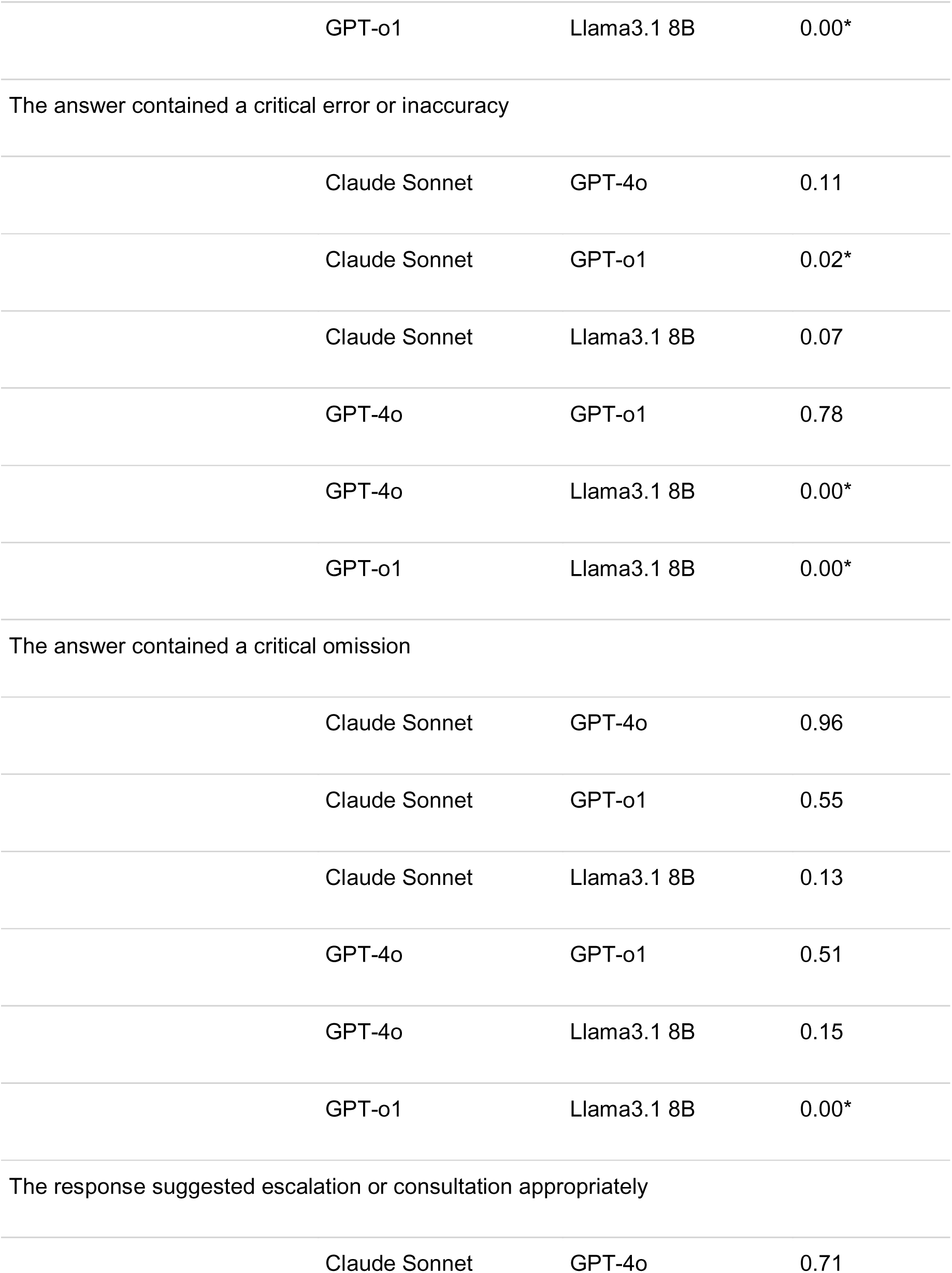
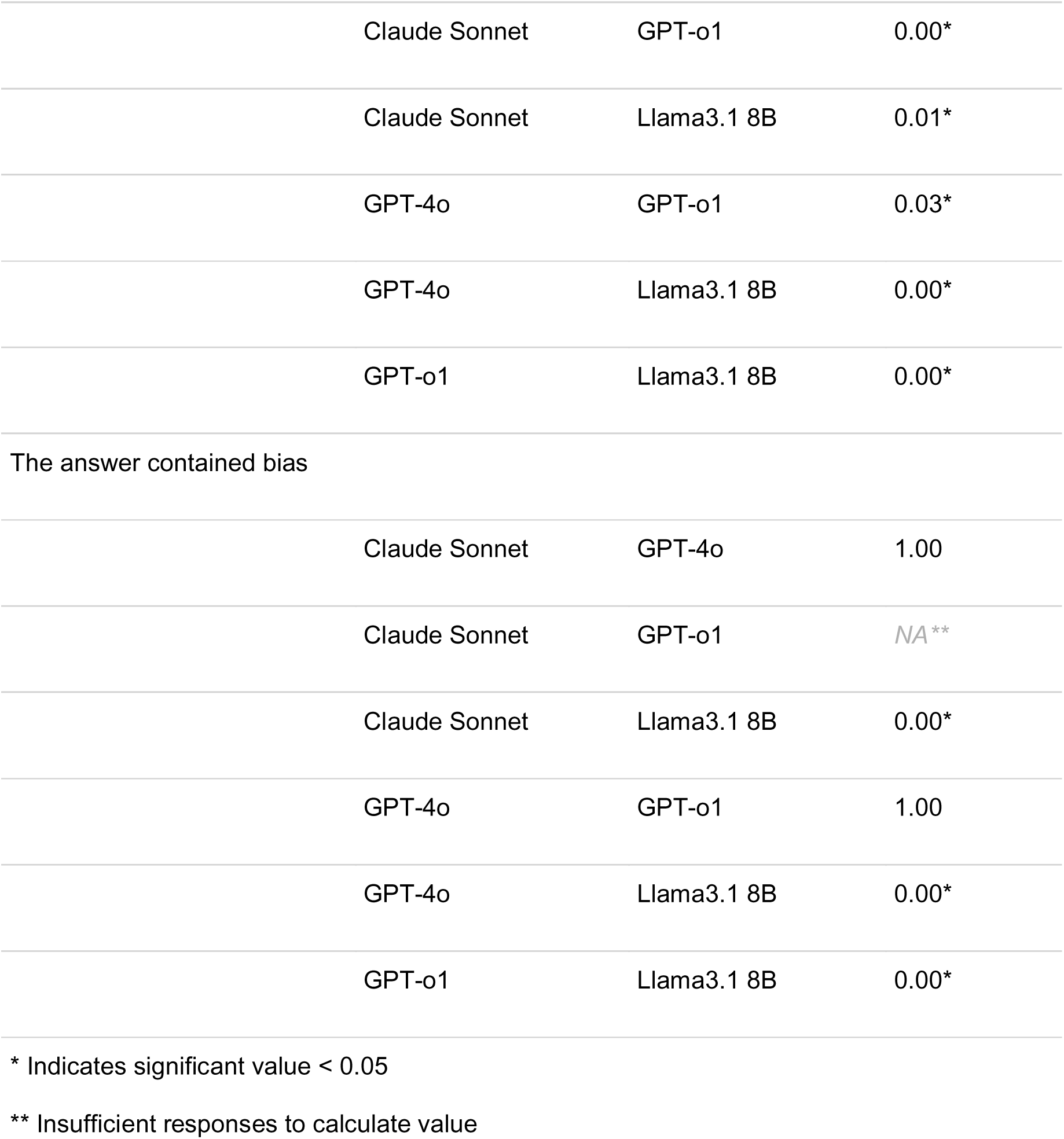
Pairwise comparison of models on general medical criteria Characteristic.

### Performance on infectious diseases specific criteria

With respect to ID-specific criteria, the other three models significantly outperformed Llama for accurate interpretation of microbiology results, adherence to AMS principles, antibiotic suggestion with appropriate spectrum, infection control consideration and appropriate involvement of other disciplines (Table 5). On accurately interpreting microbiology results, GPT-o1 scored significantly higher than GPT-4o (90 vs 57%, p=0.01) and not Claude (90 vs 67%, p=0.77). Regarding adherence to AMS principles GPT-o1 and GPT-4o significantly outperformed Claude (GPT-o1 93% vs Claude 40%, p=0.00, GPT-4o 77% vs Claude 40%, p=0.03) but were not significantly different.

**Table 5:**
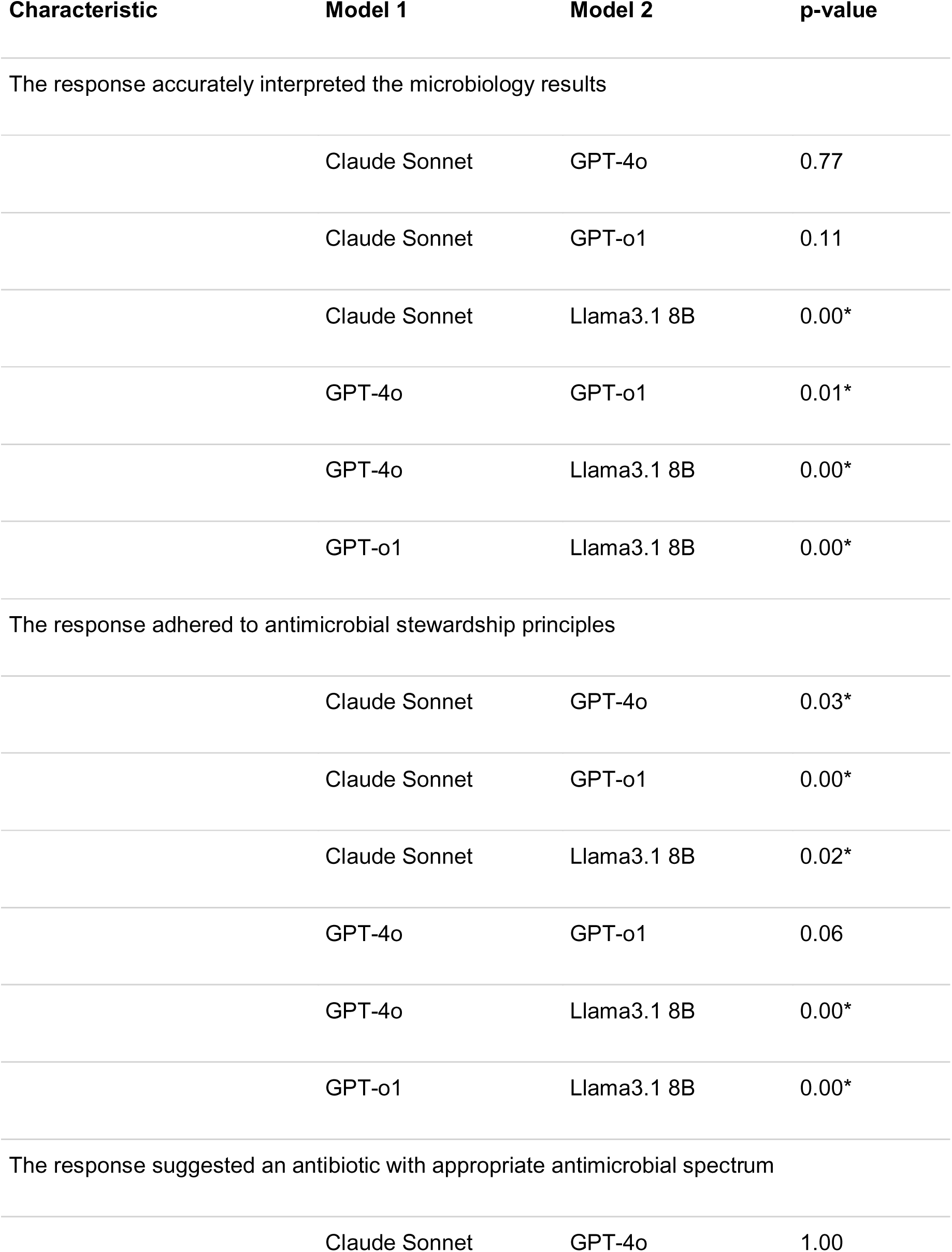
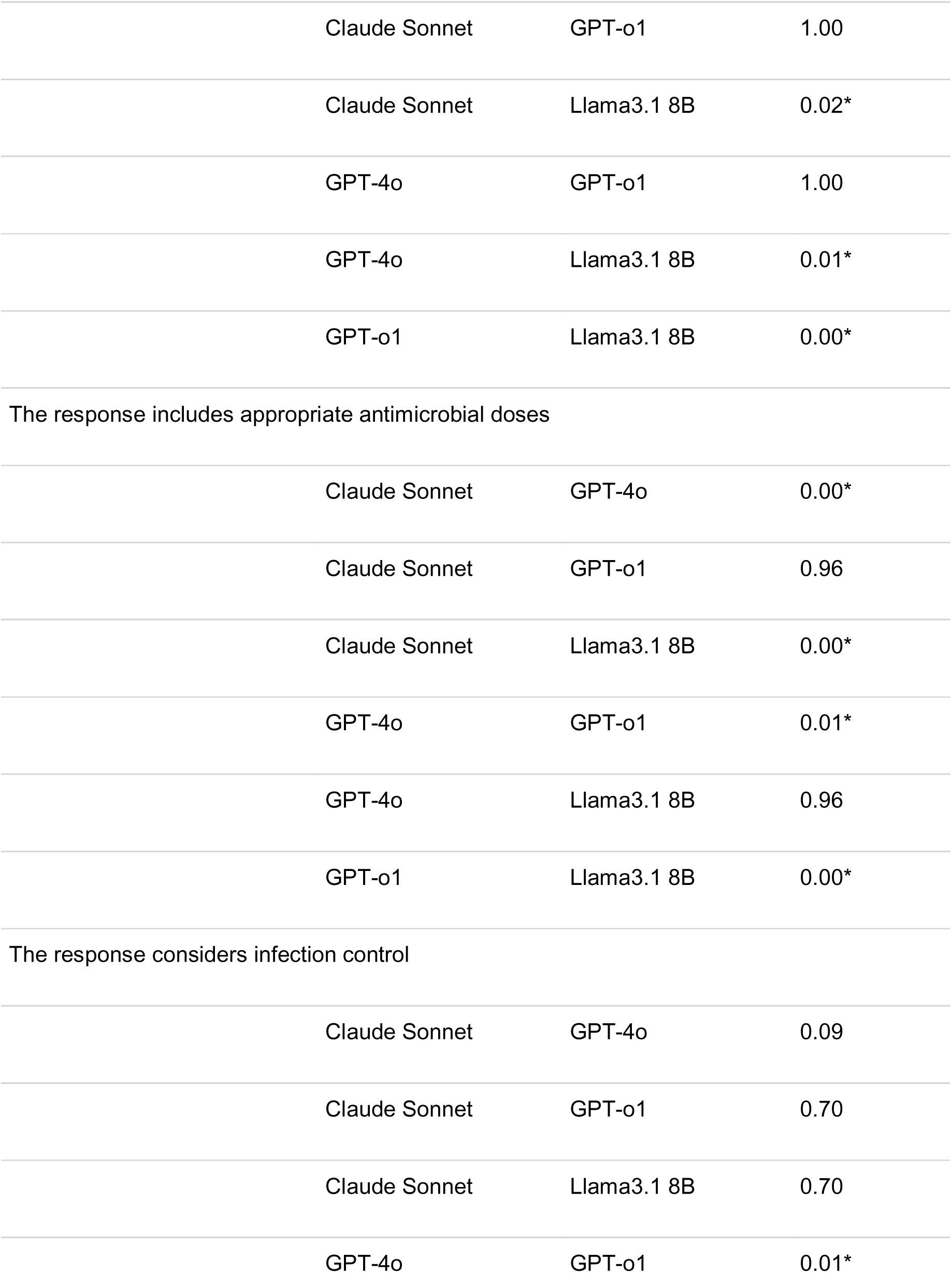
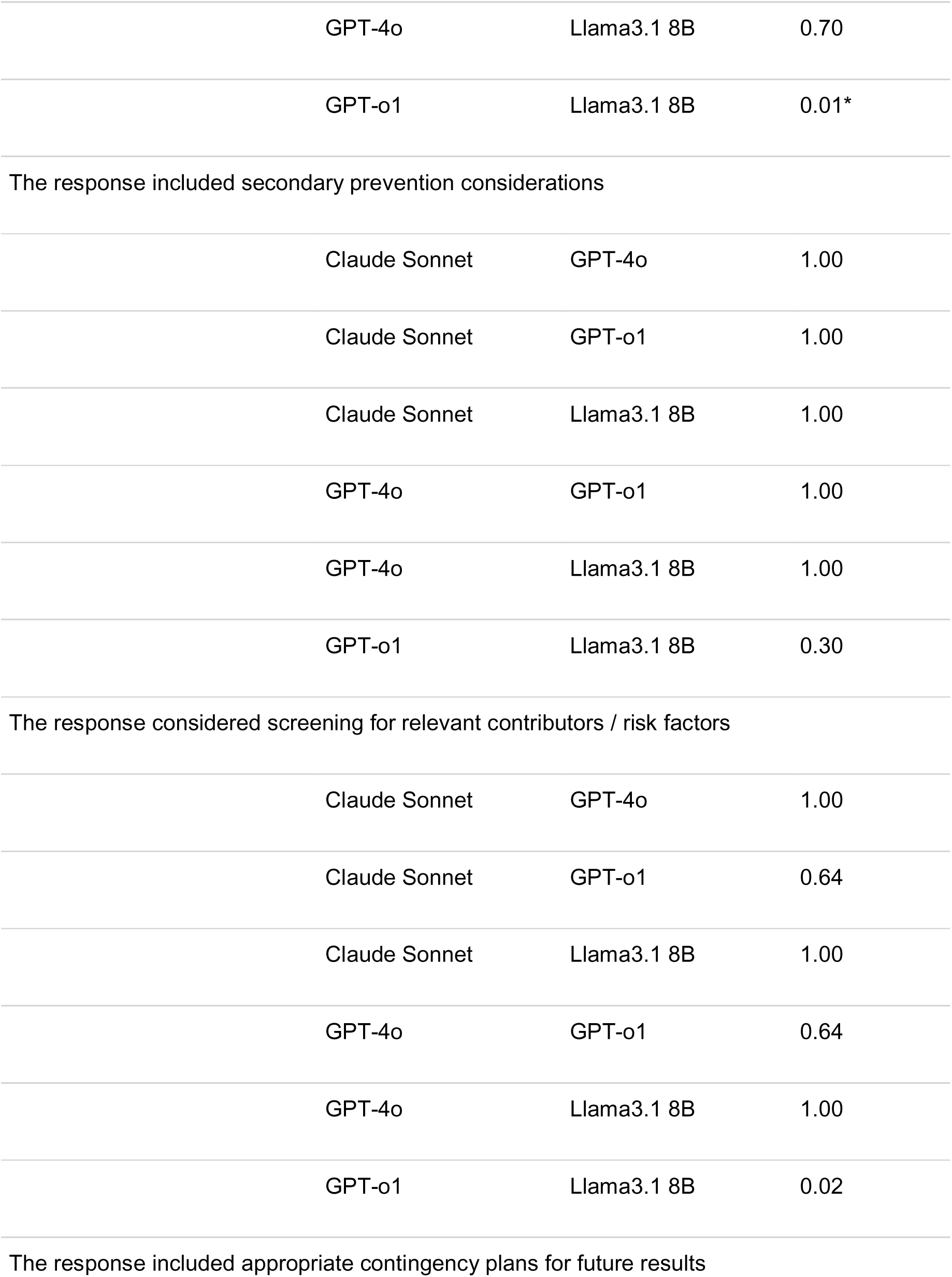
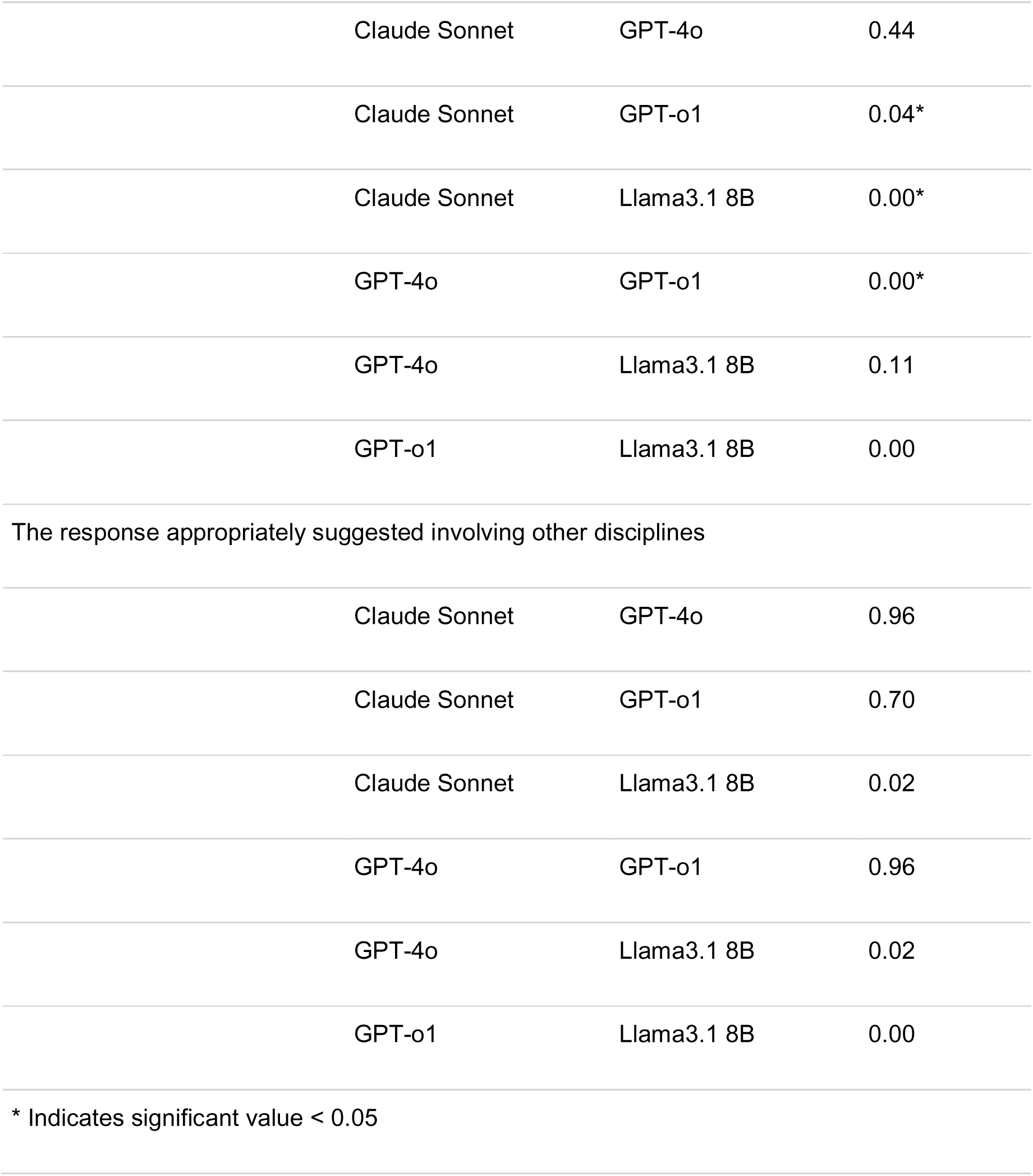
Pairwise comparison of models on infectious disease specific criteria Characteristic.

GPT-o1 and Claude both outperformed the other models on appropriate antimicrobial dosing and infection control (Table 2 and 4). GPT-o1 also outperformed all models on appropriate contingency planning for future results (GPT-o1 70% vs Claude 20%, p=0.04, GPT-4o 20% p=0.00). GPT-o1, GPT-4o and Claude did not differ significantly on the criteria of appropriate antimicrobial spectrum or involvement of other disciplines.

Llama performed poorly, interpreting microbiology accurately in 3.3% of responses, adhering to AMS principles in 10% of responses, and suggesting appropriate antimicrobial spectrum and doses in 20% and 21% of responses, respectively. All models performed similarly poorly regarding advice for secondary prevention (Claude 11%, GPT-4o 5.6%, GPT-o1 24%, Llama 5.3%, p = 0.7) and assessment and management of risk factors (percentage agreement Claude 20%, GPT-4o 20%, GPT-o1 37%, Llama 13%, p = 0.12).

### Effect of multiple reviewers, scenarios and models on performance evaluation

We performed a mixed-effect regression model to estimate the relative effects of the model, scenario, reviewer and model replicate on ratings (Table 6). Compared with the reference model (GPT-4o), GPT-o1 had 1.6-times higher odds (OR 1.62, 95% confidence interval (CI) 1.23 - 2.13) of receiving higher ratings, whereas Llama had an odds ratio of 0.39 (95% CI 0.30 - 0.52). Scenario 3 was associated with an odds ratio of 0.65 for receiving a high rating in comparison to Scenario 1 (OR 0.65, 95% CI 0.48 - 0.87). Model replicate and the reviewer were not significant, indicating that the LLM performance had the strongest impact on the rating.

**Table 6:** Ordinal link mixed model results for effect on scoring.

| Term | Odds ratio (95% CI) | p-value |
| --- | --- | --- |
| Model (vs Claude Sonnet) |  |  |
| GPT-4o | 0.93 (0.71, 1.23) | 0.61 |
| GPT-o1 | 1.62 (1.23, 2.13) | 0.0005 |
| Llama3.1 8B | 0.39 (0.30, 0.52) | < 0.001 |
| Model pass (vs. pass 1) |  |  |
| Pass 2 | 0.98 (0.79, 1.22) | 0.88 |
| Pass 3 | 0.86 (0.69, 1.07) | 0.16 |
| Reviewer B (vs. A) | 1.10 (0.93, 1.30) | 0.27 |
| Scenario (vs. 1) |  |  |
| Scenario 2 | 1.00 (0.73, 1.37) | 0.98 |
| Scenario 3 | 0.65 (0.48, 0.87) | 0.0054 |
| Scenario 4 | 1.05 (0.77, 1.44) | 0.74 |
| Scenario 5 | 0.91 (0.68, 1.23) | 0.56 |

## Discussion

Using a series of five complex ID case vignettes, we demonstrate significant variation in performance of four LLMs that vary in size and reasoning capability. The open weight Llama 3.1 8B model performed consistently poorly. The large, non-reasoning models had variable performance, and the only reasoning model, GPT-o1 had better, but inconsistent performance. Overall, our study shows that these LLMs remain suboptimal for diagnostic and management support for complex real-world cases faced by ID consultants.

The performance disparity between the models is expected, given that the locally hosted Llama 3.1 8 billion parameter model is orders of magnitude smaller than the GPT-o1, which, although not publicly disclosed, is likely trained with trillions of parameters [23]. However, we include this model for comparison, as local models can potentially address privacy concerns with cloud-based LLMs, especially for physicians that do not have institutional access to a LLM with privacy safeguards. The poor performance of Llama 3.1 8B highlights the potential performance pitfalls with local models on complex clinical reasoning tasks. Similarly, the improved performance of GPT-o1 over the non-reasoning models is expected but all three require agreements between health systems and the software developer to protect patient data, meaning these models may only be available in large, well-funded healthcare systems.

Our findings suggest that for sub-specialty management plans, LLMs do not perform at levels safe for deployment. In this study, LLM performance across general medical criteria was worse than previously reported performance in complex diagnostics cases [24] and script concordance tests [25] where reasoning models typically had a 50-65% accuracy on stepwise diagnosis and reasoning across multiple specialties. The assessment that 30 to 40% of responses from the non-local models were unsafe and 30 to 53% contained a critical error is likely because, unlike previous assessments that focus largely on diagnosis and management of common conditions encountered by generalists, these scenarios focused on complex, sub-specialty management. This finding is corroborated by recent evidence regarding the use of OpenEvidence, a RAG-based LLM resource, where performance on sub-specialty medical questions demonstrated 34 to 41% accuracy, even with reasoning [26].

When the task was specifically outlined in the prompt, for example including antibiotic dosing, the models performed well on ID-specific criteria. However, when ID specific considerations such as assessment of risk factors or secondary prevention were not explicitly included, they were ignored by all models. This oversight highlights a potential risk of LLM use for advice by the non-specialist, as, if the ID implications are not considered and explicitly sought from the LLM, they are likely to be omitted from the plan.

This study has several strengths, including evaluation of management plans for complex ID cases as opposed to multiple choice questions datasets or routine cases. Our study has several weaknesses inherent to many current studies on LLMs, in that the models assessed have since been superseded by more sophisticated versions. The most impactful omission is of more recent open-source reasoning models such as DeepSeek, Qwen and gpt-oss-120b [27]. Additionally, we assessed “out-of-the-box” models that are not fine-tuned for medical tasks or augmented by RAG or agentic AI workflows. Recent evidence suggests that models with RAG such as OpenEvidence outperformed GPT-4o and Gemini 2.5 Pro when applying guidelines [12]. However, this performance cannot necessarily be extrapolated to our data given these cases are not covered by routine guidelines. Similarly, single or multi-agent models that may seek out further information and iterate over responses may increase performance but are yet to be routinely implemented into practice settings [28]. We also did not give the models the information sequentially as would occur in medical practice, but in one zero-shot prompt, which may reduce generalizability to real-world settings. Despite these limitations, we feel our methodology reflects the way in which most clinicians without specific AI expertise or training are likely to access and use LLMs.

LLMs are playing an increasingly significant role in clinical care. This pragmatic assessment of commonly available models showed that current performance varies when providing ID advice in complex scenarios. All but the most sophisticated LLM gave treatment plans with a risk of harm or critical omission in more than 30% of cases. Moreover, performance on specialty-specific considerations such as AMS and secondary prevention varied substantially between models. This study highlights the need for caution in application of out-of-the-box LLMs to complex ID workflows that reflect real-world practice.

## Data Availability

All data produced in the present study are available upon reasonable request to the authors

## Supplementary Materials

### Supplementary 1: Scenario example

#### Prompt

You are an infectious diseases attending. Please give a management plan for the following consult including medication dosing, antimicrobial stewardship, infection control as relevant. Ms. Supatra is a 25-year-old female with no known past medical history, who is admitted under the neurology service with over a week of increasing confusion and headaches. On initial examination she had a temperature of 38.1°C, was disorientated, with a negative Kernig’s sign and no nuchal rigidity. She had increased tone in her lower limbs but normal power. Fundoscopic examination does not reveal papilledema.

An initial lumbar puncture performed in the emergency room demonstrated the following cerebrospinal fluid (CSF) profile:

|  |  |
| --- | --- |
| Opening pressure | 160 mm H<sub>2</sub>O |
| Protein | 150 mg/dL |
| Glucose | 80 mg/dL (serum glucose 90 mg/dL) |
| WBC | 35 WBC/mm<sup>3</sup> with a lymphocytic predominance |
| Gram stain | Negative |

She is started empirically on intravenous ceftriaxone, ampicillin and acyclovir but she shows no improvement in her cognition over the next 4 days. A lumbar puncture is repeated and demonstrates the following:

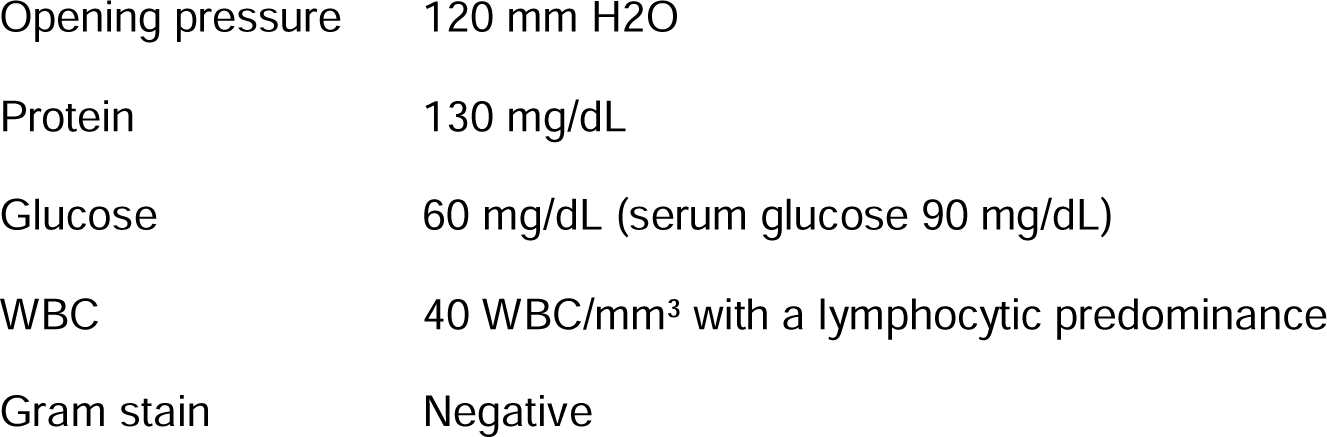

Other microbiology results including CSF culture, multiplex PCR, cryptococcal antigen on blood and CSF and HIV serology are negative to date. An interferon gamma release assay for latent tuberculosis is negative. Computer tomography of her brain on admission does not show any abnormalities and an MRI will occur in the coming days. Autoimmune encephalitis antibodies are pending.

Ms Supatra was born in rural Thailand and migrated to the US at age 19. She has returned twice since then. Her family denies any preceding infectious symptoms.

### Supplementary Material 2: Evaluation metrics

Evaluation Questions - General

1. The model correctly interpreted the scenario
2. The response was accurate
3. The response was complete / comprehensive
4. The communication in the response was clear
5. The response was unsafe (ie. there was a risk of harm from the response)
6. The answer contained a critical error or inaccuracy
7. The answer contained a critical omission
8. The response suggested escalation or consultation appropriately
9. The response contained bias

Evaluation Questions - ID-specific evaluation

1. The response accurately interpreted the microbiology results
2. The response adhered to antimicrobial stewardship principles (narrow spectrum, short duration, antibiotic rationalization)
3. The response suggested an antibiotic with appropriate antimicrobial spectrum
4. The response includes appropriate doses of antimicrobials
5. The response considered infection control
6. The response included secondary prevention considerations (ie. MRSA decolonization, vaccination, latent TB therapy)
7. The response considered screening for relevant contributors / risk factors (ie. immunosuppression, IDU)
8. The response included appropriate contingency plans for future results (ie. If no cultures return in X time then do Y action)?
9. The response suggested involving other disciplines / specialties as appropriate

**Supplemental Table 1:**
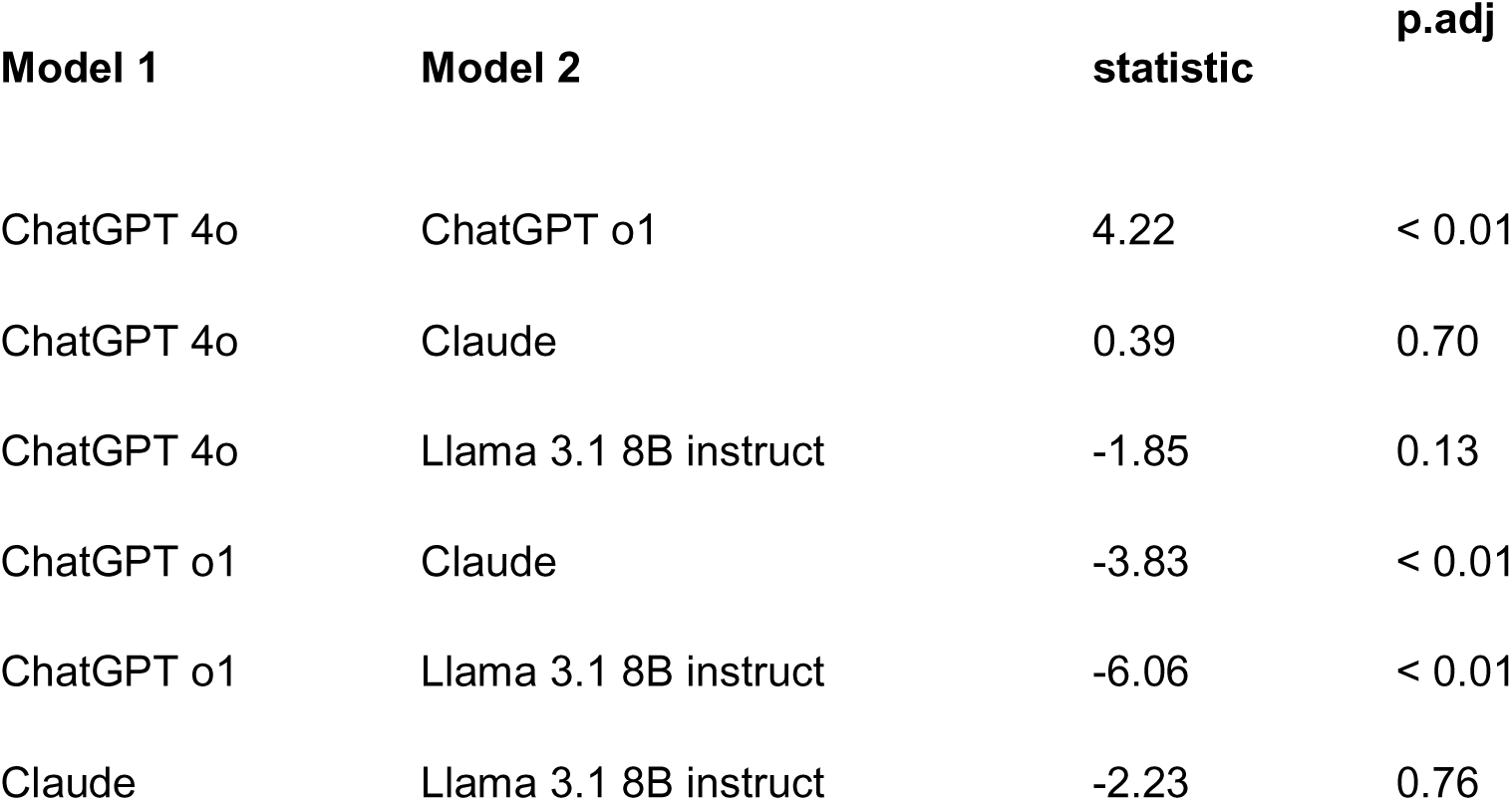
Pairwise comparison of model response lengths using Dunn test.

