## Supplement for "Evaluation of four large language models on complex, infectious disease case scenarios"

**Supplementary Materials**

**Supplementary 1: Scenario example**

**Prompt:**

You are an infectious diseases attending. Please give a management plan for the following consult including medication dosing, antimicrobial stewardship, infection control as relevant.

Ms. Supatra is a 25-year-old female with no known past medical history, who is admitted under the neurology service with over a week of increasing confusion and headaches. On initial examination she had a temperature of 38.1°C, was disorientated, with a negative Kernig’s sign and no nuchal rigidity. She had increased tone in her lower limbs but normal power. Fundoscopic examination does not reveal papilledema.

An initial lumbar puncture performed in the emergency room demonstrated the following cerebrospinal fluid (CSF) profile:

Opening pressure	160 mm H2O

Protein	150 mg/dL

Glucose	80 mg/dL (serum glucose 90 mg/dL)

WBC	35 WBC/mm³ with a lymphocytic predominance

Gram stain	Negative

She is started empirically on intravenous ceftriaxone, ampicillin and acyclovir but she shows no improvement in her cognition over the next 4 days. A lumbar puncture is repeated and demonstrates the following:

Opening pressure	120 mm H2O

Protein	130 mg/dL

Glucose	60 mg/dL (serum glucose 90 mg/dL)

WBC	40 WBC/mm³ with a lymphocytic predominance

Gram stain	Negative

Other microbiology results including CSF culture, multiplex PCR, cryptococcal antigen on blood and CSF and HIV serology are negative to date. An interferon gamma release assay for latent tuberculosis is negative. Computer tomography of her brain on admission does not show any abnormalities and an MRI will occur in the coming days. Autoimmune encephalitis antibodies are pending.

Ms Supatra was born in rural Thailand and migrated to the US at age 19. She has returned twice since then. Her family denies any preceding infectious symptoms.

**Supplementary Material 2: Evaluation metrics**

Evaluation Questions - General

1. The model correctly interpreted the scenario
2. The response was accurate
3. The response was complete / comprehensive
4. The communication in the response was clear
5. The response was unsafe (ie. there was a risk of harm from the response)
6. The answer contained a critical error or inaccuracy
7. The answer contained a critical omission
8. The response suggested escalation or consultation appropriately
9. The response contained bias

Evaluation Questions - ID-specific evaluation

1. The response accurately interpreted the microbiology results
2. The response adhered to antimicrobial stewardship principles (narrow spectrum, short duration, antibiotic rationalization)
3. The response suggested an antibiotic with appropriate antimicrobial spectrum
4. The response includes appropriate doses of antimicrobials
5. The response considered infection control
6. The response included secondary prevention considerations (ie. MRSA decolonization, vaccination, latent TB therapy)
7. The response considered screening for relevant contributors / risk factors (ie. immunosuppression, IDU)
8. The response included appropriate contingency plans for future results (ie. If no cultures return in X time then do Y action)?
9. The response suggested involving other disciplines / specialties as appropriate

Supplemental Table 1: Pairwise comparison of model response lengths using Dunn test

| **Model 1** | **Model 2** | **statistic** | **p.adj** |
| --- | --- | --- | --- |
| ChatGPT 4o | ChatGPT o1 | 4.22 | < 0.01 |
| ChatGPT 4o | Claude | 0.39 | 0.70 |
| ChatGPT 4o | Llama 3.1 8B instruct | -1.85 | 0.13 |
| ChatGPT o1 | Claude | -3.83 | < 0.01 |
| ChatGPT o1 | Llama 3.1 8B instruct | -6.06 | < 0.01 |
| Claude | Llama 3.1 8B instruct | -2.23 | 0.76 |
